# Two Color Single Molecule Sequencing on GenoCare™ 1600 Platform to Facilitate Clinical Applications

**DOI:** 10.1101/2020.09.28.20203455

**Authors:** Fang Chen, Bin Liu, Meirong Chen, Zefei Jiang, Zhiliang Zhou, Ping Wu, Meng Zhang, Huan Jin, Linsen Li, Liuyan Lu, Qi Wang, Huan Shang, Bing Xie, Lei Liu, Xia Lin, Weiyue Chen, Jianfeng Xu, Ruitao Sun, Guangming Wang, Jiao Zheng, Jifang Qi, Bo Yang, Dexia Chen, Lidong Zeng, Gailing Li, Yan Li, Hui Lv, Nannan Zhao, Bo Zhou, Wen Wang, Jinsen Cai, Siyu Liu, Weiwei Luo, Juan Zhang, Yanhua Zhang, Yongyi Lu, Jicai Fan, Haitao Dan, Xuesen He, Lichun Liu, Yan Feng, Jianglin Chen, Wei Huang, Lei Sun, Qin Yan

## Abstract

With the rapid development of precision medicine industry, DNA sequencing becomes increasingly important as a research and diagnosis tool. For clinical applications, medical professionals require a platform which is fast, easy to use, and presents clear information relevant to definitive diagnosis. We have developed a single molecule desktop sequencing platform, GenoCare™ 1600. Fast library preparation (without amplification) and simple instrument operation make it friendlier for clinical use. Here we presented sequencing data of E. coli sample from GenoCare™ 1600 with consensus accuracy reaches 99.99%. We also demonstrated sequencing of microbial mixtures and COVID-19 samples from throat swabs. Our data show accurate quantitation of microbial, sensitive identification of SARS-CoV-2 virus and detection of variants confirmed by Sanger sequencing.

## INTRODUCTION

Starting from 1990, National Human Genome Research Institute (NHGRI) spent 14 years assembling the first version of the human genome [1]. During and after that decade, different sequencing techniques have been developed and commercialized [2-9]. As a result, high throughput, low-cost genomic sequencing techniques revolutionized biomedical research, clinical diagnosis and made it one important foundation for precision medicine [10-13]. Among those techniques, single molecule (SM) sequencing technologies are well known as amplification-free sequencing, which avoid the errors and biases from clonal amplification involved in NGS platforms [14].

Based on different sequencing principles, there are three categories of SM techniques with unique characteristics reported over the past decades: (1) sequencing-by-synthesis (SBS) based true single molecule sequencing (tSMS) from Helicos Biosciences [5, 15]; (2) single molecule real time (SMRT) from Pacific Biosciences, which utilizes “zero-mode waveguide” to monitor fluorescence signals emitted of single DNA polymerase extension [9, 16]; (3) nanopore sequencing from Oxford Nanopore Technologies (ONT), which measures the current change when DNA translocates through protein pore [8, 17]. Although SMRT and nanopore approaches could generate long read length (>10kbp), their applications are limited by low number of reads, high raw error rate and expensive sequencing flow cells.

We have developed an SBS based single molecule sequencing platform, which provides enough reads throughput and accuracy with the advantages of being amplification-free. Compared to tSMS from Helicos, our GenoCare™ 1600 desktop platform is based on two color chemistry, which improves real reaction efficiency and reduces the sequencing time. Aiming at clinical applications, GenoCare™ 1600 is optimized on library preparation, flow cell design, instrument stability, basecalling algorithm and bioinformatics software, etc. In this paper, we used GenoCare™ 1600 to sequence E. coli sample, microbial mixtures, and COVID-19 patient samples from throat swab. Within 15 hrs, we got E. coli sequencing reads with average read length of 53 bp, and consensus accuracy exceeding 99.99%. Microbial mixture sequencing results show quantitative response of sample concentrations. The identification of positive COVID-19 and detection of variants demonstrated its possibilities in clinical applications.

## RESULTS AND DISCUSSION

### Platform and workflow of two-color single molecule sequencing

#### Two color single molecule sequencing chemistry

We have developed four reversible terminators with 3’-OH unblocked and inhibition structure on the side chain extended from the base ring (Figure **1A**) for sequencing by SBS on single molecule level. Two terminators are labeled with a green dye, whose peak fluorescent emission wavelength is 552 nm, while the other two terminators are labeled with a red dye with peak fluorescent emission at 664 nm. For SBS technique, polymerase inhibition is one important parameter for terminator nucleotides. To test the terminator’s inhibition performance, we developed a solution reaction method with different types of oligo templates. When oligo templates are hybridized with FAM-labeled primers, terminators, polymerase and reaction buffers are added and the terminators are incorporated to the end of primers. After a full reaction, the reaction product is analyzed using capillary electrophoreses (ABI 3100), which shows one-base resolution. Controlling reaction conditions, we first ensure only one terminator (Adenosine) is added to the primer (Figure **1B, 1C**). Due to the side chain modification structure on the terminator, with this one terminator addition, the primer’s mobility will move from 53 to more than 60. The same reaction has been conducted using templates with triple T, and the results indicated only one terminator incorporation (Figure **1D**). In addition, we introduced both terminators A and T to reaction system with template sequence of TA. After reaction, the same result is shown (Figure **1E**). With this solution reaction model, we designed and confirmed a series of structures’ inhibition performance and selected the best structures for each terminator.

**Figure 1:**
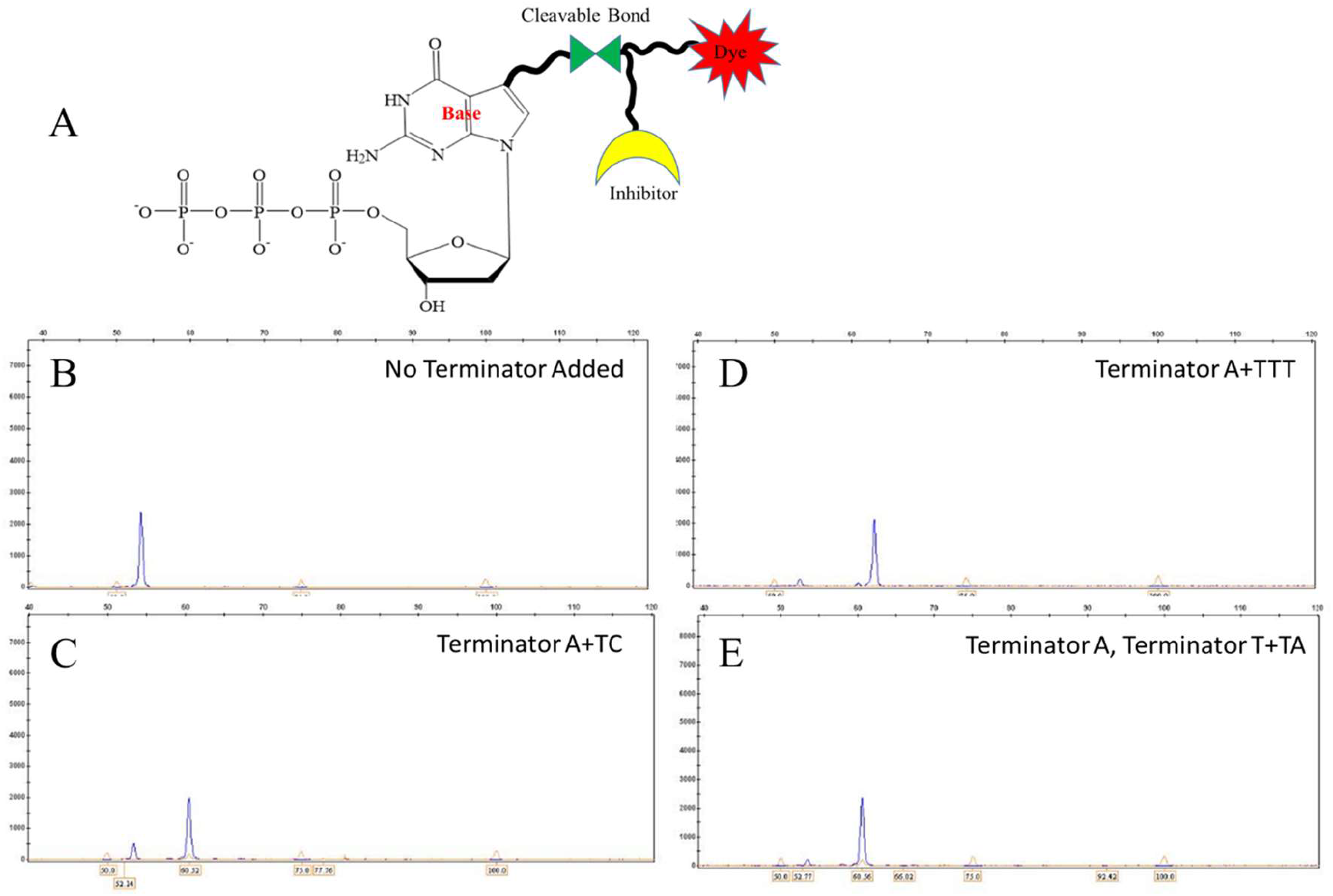
**A**. The scheme of terminator structure; **B-E**: capillary electrophoresis analysis result for different reactions (reaction conditions shown in figure).

The other key parameter to consider is fluorescence intensity of the labels. To optimize the label structure, the properties of dyes, including photo-stability, hydrophobicity, and quantum yield, etc. are evaluated [18]. In addition, considering the redox properties of four nucleotide bases [19], for example, photoinduced electron transfer (PET) effect of G [20], which decreases the fluorescent signal intensities, the cleavable linker in Figure **1A** is designed to mitigate the bases’ quenching effect. With optimized terminator structures, the consistently high signal-to-noise ratio (Supplementary Information: **S1**) could be assured in sequencing.

We also modified the polymerase to improve properties such as thermo-stability, extension rate, fidelity, specificity, damage bypass, inhibitor resistance, strand displacement, and especially, the capability to incorporate terminators. Initially, the wild-type Klenow fragment with Mn2+ as divalent cation was applied with our terminators. However, Mn2+ could cause high probability of mis-incorporation, which will lead to high error rate or even complete termination due to the mismatch event. Therefore, we engineered and selected the polymerases which could catalyze our terminator reaction with Mg2+ with high efficiency and fidelity.

During each sequencing cycle, two terminators with different colors are added and incorporated to primers on surface. The inhibitor structure ensures single terminator complementary to template extension. After washing off un-reacted terminators, surface images are captured under the protection of imaging buffer. Then the inhibitor structure and dye are cleaved to resume reaction for the next cycle (Figure **2**), in which the other two terminators are incorporated. According to different applications, different number of sequencing cycles are selected to balance the total reaction time, reagent cost and read length. Compared to NGS systems, single molecule sequencing obliviates the pre-phasing and phasing problem caused by the asynchronous reaction in amplified cluster, which complicates NGS sequencing chemistry and data analysis.

**Figure 2.**
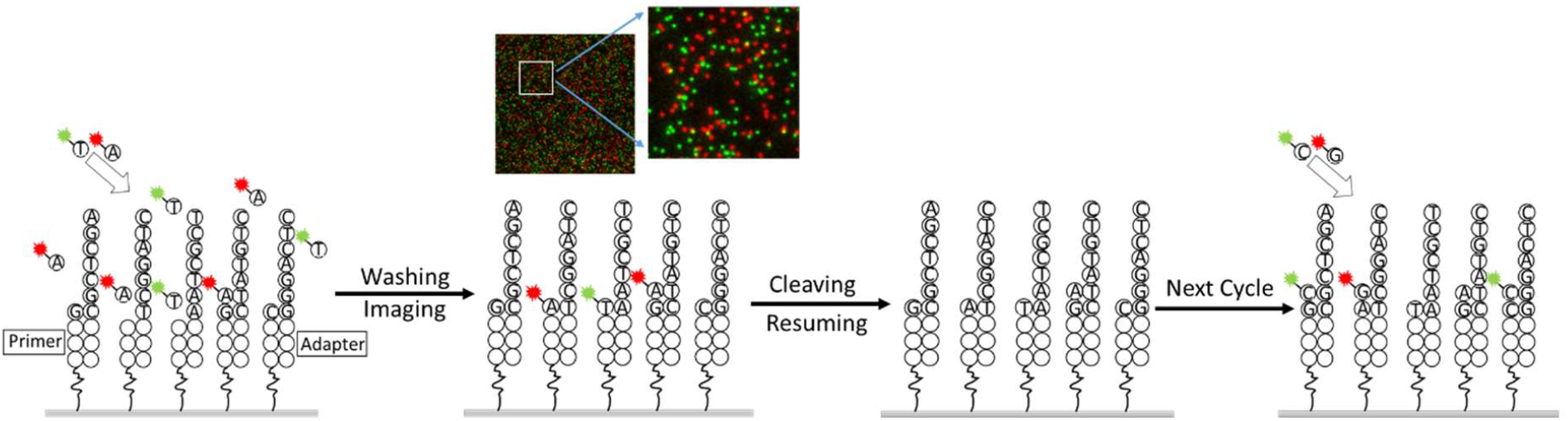
The scheme of sequencing cycle.

#### Library preparation

One major advantage for single molecule sequencing over amplification-based NGS technology is that we can develop much simpler library preparation process without the cluster generation step. Figure 3 illustrates the general workflows for GenoCare™’s library preparation, which cost only 1.5 hrs and 2.8 hrs for DNA and RNA samples respectively.

**Figure 3.**
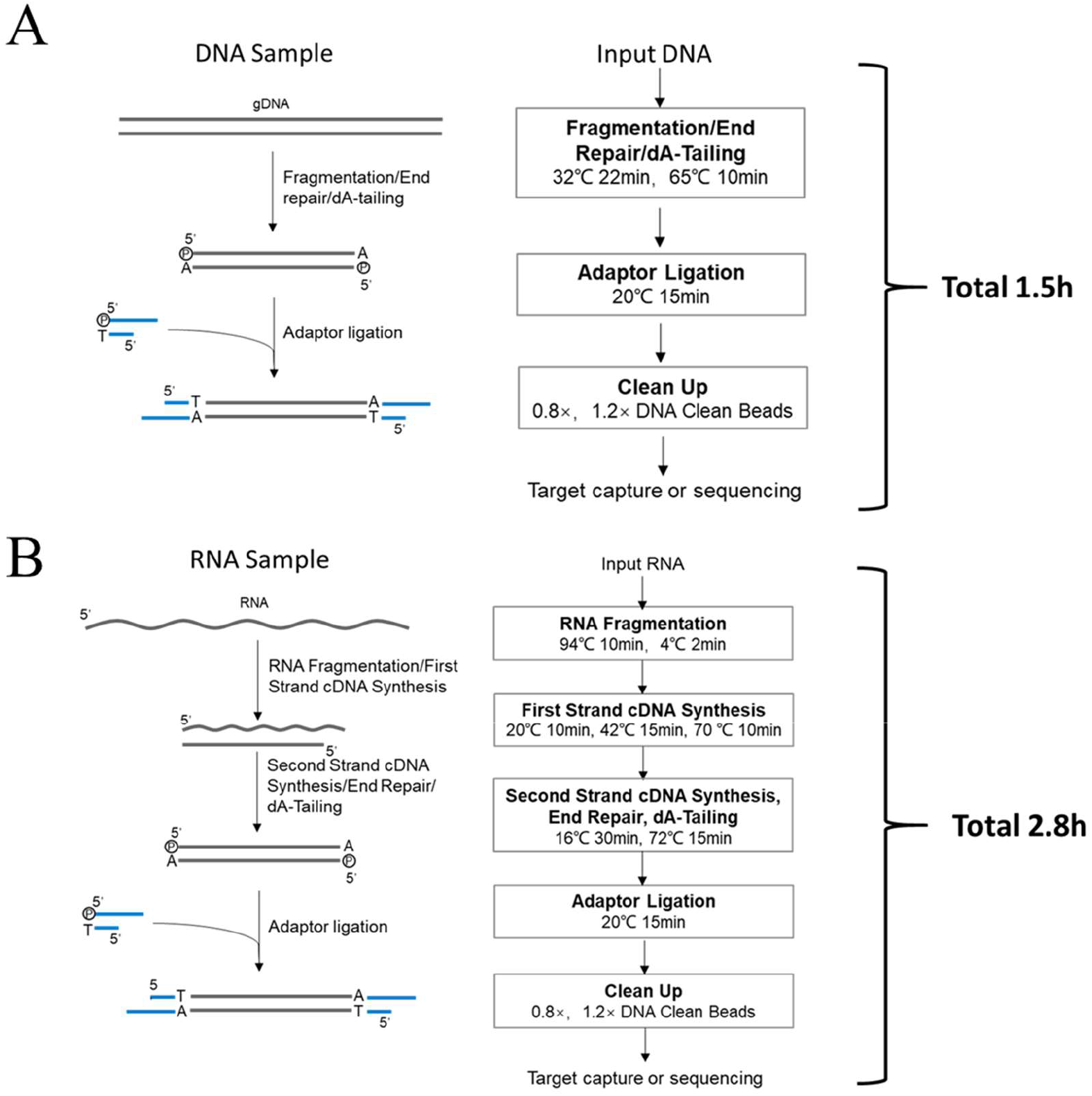
Universal workflow of library preparation for single molecule sequencing: (A) DNA sample; (B) RNA sample.

A straightforward benefit from GenoCare™’s library preparation is that the libraries with single-end ligated adaptors are also applicable for sequencing. Compared to traditional NGS platforms which only amplify libraries with double-end ligated adaptors, Genocare™ uses samples more efficiently, which makes it particularly suitable for minimal amount samples such as cfDNA. To be specific, our library preparation can start from as low as 3ng of DNA.

#### Surface hybridization

The surface chemistry of GeneMind’s flow cell was described in previous publications [21, 22]. Briefly, primers immobilized on flow cell surface are designed to capture the samples with sequencing adaptors. In GenoCare™ 1600 system, a 16-lane flow cell is assembled with a functionalized glass coverslip (110 mm*74 mm) and a bottom glass slide using pressure sensitive adhesive. To maximize the uniformity and single molecule ratio, we first coat coverslip with single layer of epoxy silane using the chemical vapor deposition (CVD) technology. A 62nt-long oligo with -NH2 modification at 5’ end is designed and anchored on surface as a universal probe (the primer could be customized upon requirements of specific applications). The probe contains ∼50% of GC, which specifically hybridizes sample DNAs and stabilize them during sequencing process. The library’s adaptor is modified with extra bases at 3’ position to prevent extension from library strand’s 3’ end. Compared to poly-T capture probe utilized in previous reports, this new design eliminates the fill-lock step and reduces total sample loading time to 30 min. It also enables direct capture of preselected region of genome in targeted gene sequencing.

#### Sequencer hardware

GenoCare™ 1600 (GeneMind, Shenzhen, China) is designed as a desktop platform for two color single molecule sequencing. The instrument adopts wide field total-internal-reflection-fluorescence (TIRF) optics to detect the weak signals from single molecule nucleotides. In the TIRF system, green and red lasers with emitting wavelengths at 532 nm and 639 nm are coupled together to simultaneously illuminate the samples through the edge of a high NA objective. The illumination generates an evanescent light wave layer as shallow as 100 nm beyond the glass-solution interface, which can excite the fluorescence labels of terminators on surface while leave the vast majority of the background un-illuminated. To maximize the signal-to-noise ratio (SNR) of single molecules, the imaging system utilizes two scientific CMOS cameras with both high quantum efficiency and low noise to photograph each field of view (FOV).

Image stabilization is a tough challenge for high-sensitivity single molecule optics like TIRF. Single molecule image quality is vulnerable to multiple sources including but not limited to environmental vibration, stage motion, temperature fluctuation and air disturbance. As the depth-of-field of the TIRF objective is as small as 500 nm, a high-accuracy auto-focusing system is installed to lock the imaging focus in real time during XY scanning of the flow cell. Besides, multiple-level anti-vibration design is performed on the mechanical structure of the instrument to minimize the influence from external and internal vibration sources. To speed up the sequencing process, an entire flow cell is virtually divided into two parallel units with eight lanes each. During each sequencing cycle, when one unit is on the process of fluidic motion and biochemical reaction, the other is on imaging, and vice versa.

#### Data processing

The raw sequencing image files are analyzed in real time by the machine-learning algorithm based software DirectCall, developed by GeneMind. Generally, the workflow of the data processing includes background elimination, spot localization, image registration, template building, and basecalling. To improve mapping rate and save mapping time, a Q-score system is developed to filter the raw data from basecalling. Different from the Q-score system widely adopted by NGS platforms, GenoCare™’s Q-score is based on the unique properties of single molecule sequencing and reflects the read quality co-defined by signal intensity, SNR, template distance, etc. Higher Q-score DNA reads would probably show lower error rate and higher mapping possibility. According to applications, different Q-score is applied to filter raw data.

### Characterization of single molecule sequencing

#### Characterization of instrument performance

To characterize our sequencing system, we hybridized E. coli DNA (ATCC8739 from Guangdong Microbial Culture Collection Center) in one lane and sequenced 120 cycles with 400 FOVs’ data (That is 2.1% of recommended maximum imaging area of a flow cell) captured from each cycle (Figure **4**). Before mapping, the raw data is filtered with read length (>20bp) and Q-score. The mode peak read length is ∼70 bp and the average read length is ∼53 bp. The unique mapped reads number is 12.2M with raw mismatch rate of 0.61%, insertion rate of 1.45% and deletion rate of 2.76%. The sequencing depth is ∼130 fold, with genome coverage of 99.94% and the consensus accuracy is 99.99%.

**Figure 4.**
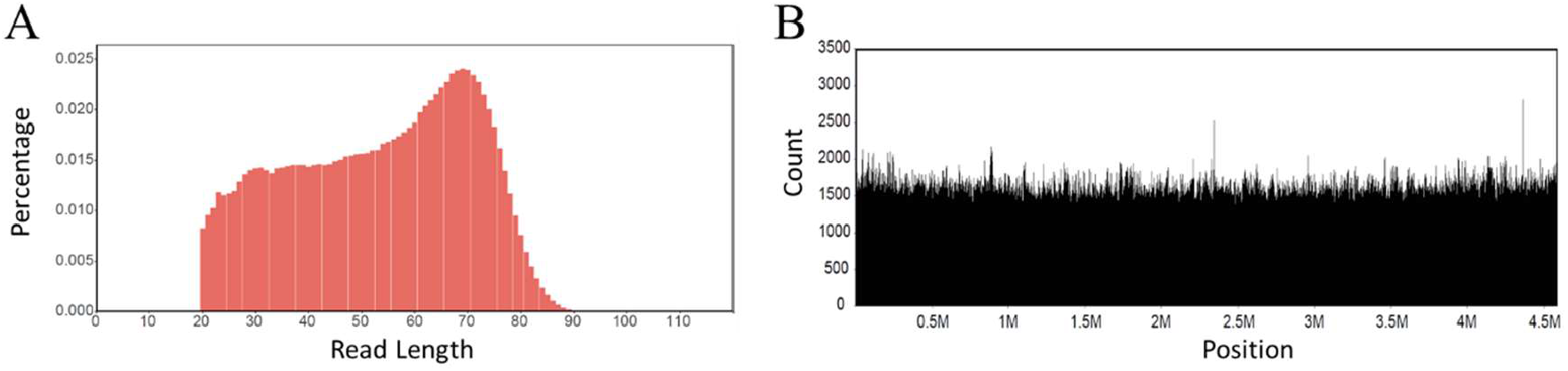
Sequencing of E. coli sample: (A) Read length distribution of unique mapping data; (B) Read counts distribution across the E. coli genome (bin size is 500 bp).

In a separate experiment, we took images from 16 lanes with 500 FOVs in each lane, and sequenced 72 cycles. Finishing within 24 hrs, 334.3M mappable reads from 16 lanes have been acquired (results shown in Supplementary Information: **S2**). The average unique mapped reads of each lane is 20.9M with CV of 4.5% ° According to genome size and bioinformatics requirement, one can optimize number of cycles, FOVs and sequencing time for best results.

#### Detection of coronavirus

In 2020, the outbreak of coronavirus (SARS-CoV-2) induced world-wide pandemic. To test the GenoCare™’s capability of sequencing microbials, we first mix Staphylococcus aureus, M13, SARS-CoV-2 (from constructed recombinant plasmid) and human DNA samples at different ratios. When acquiring 6M unique reads, the detected unique reads ratio of M13/S. aureus and SARS-CoV-2/S.aureus (log scale) have been calculated in Figure **5**. In this experiment, with changing sample ratio (M13/S. aureus, SARS-CoV-2/S.aureus) from 5e-6 to 2, the test values agree with the concentration in the sample mixtures. When acquiring 6M mapping data, the detection limit of S. aureus, M13, and SARS-CoV-2 are 10 ppm, 0.5 ppm, and 0.6 ppm, respectively. This model test indicates GenoCare™ 1600 could quantitatively identify microbials and achieve high sensitivity.

**Figure 5.**
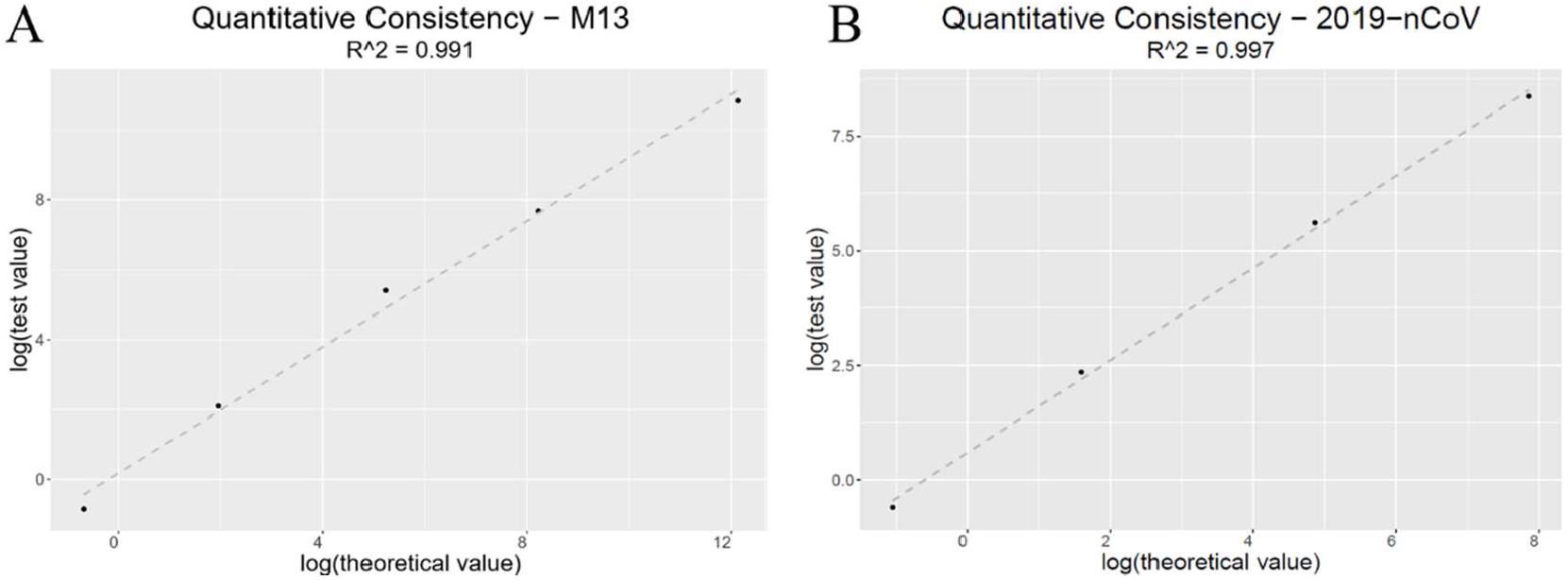
Test reads ratio vs. experimental mix ratio (log scale): (A) S. aureus/M13; (B) S. aureus/ SARS-Cov-2.

We collaborated with Shenzhen Uni-medica Co., Ltd. and acquired 3 positive samples (∼4ng/uL) from COVID-19 patients’ throat swab and a few negative samples. CDNA library was prepared by reverse-transcription, and sequencing adaptors were connected to cDNA fragments using transposase method (GenoCare DirectPrep RNA Library Prep Kit, GeneMind, Shenzhen, China. Procedure shown in Supplementary Information: **S3**). After sequencing on GenoCare™ 1600, the mapped reads over SARS-CoV-2 genome have been shown in Figure **6**. Comparing to negative samples, positive samples contain large numbers of reads which could be uniquely mapped to genome of SARS-CoV-2 reported in NCBI database. Besides, with more than 50k of reads mapped to the virus, we could see the unique mapped reads covering more than 99.9% of genome (Figure **6**). Those mismatch positions with sequencing depth more than 90x shows consistent results after resequencing using Sanger sequencing (**Table I** and Supplementary Information: **S4**), and have been proved to be mutations reported previously [23]. For examples, we detected T-C mutation on site 28144 in gene ORF 8, which shows 35.98% population frequency in *National Genomics Data Center* in February [24]. The results demonstrate the abilities of our platform in identifying and monitoring pathogenesis infections.

**Table 1.** Detected Mutations with Sanger Sequencing Verifications

| Sample ID | Position | Mutation | Depth | Sanger Sequencing |
| --- | --- | --- | --- | --- |
| 1022T | 1397 | G→A | 969 | A |
|  | 11083 | G→T | 129 | A (reverse) |
|  | 28688 | T→C | 1118 | G (reverse ) |
|  | 29742 | G→T | 294 | T |
|  | 29776 | A→T | 95 | T |
| 1028T | 8782 | C→T | 290 | A (reverse) |
|  | 8937 | C→T | 167 | A (reverse) |
|  | 28144 | T→C | 231 | C |
|  | 28878 | G→A | 239 | A |
|  | 29742 | G→A | 184 | A |

**Figure 6.**
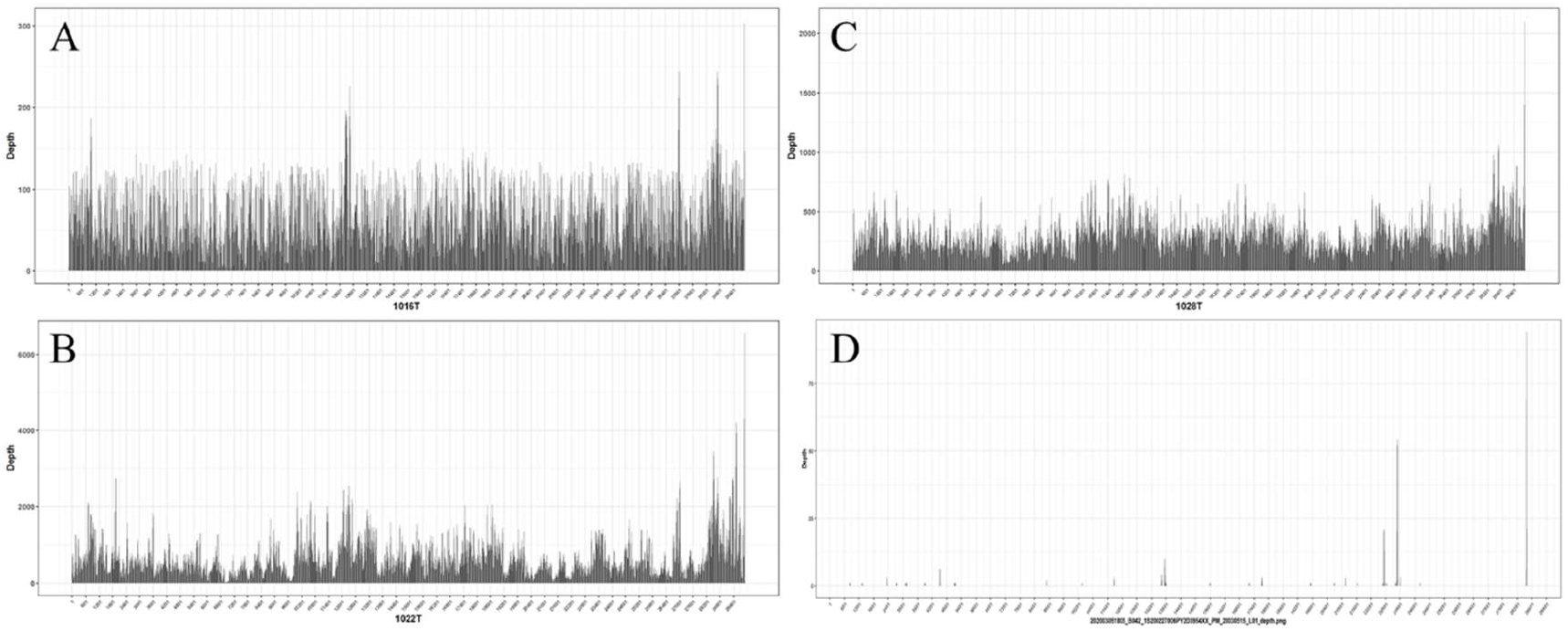
Mapping reads of positive (A, B, C) and negative (D) samples across SARS-CoV-2 genome. (The total unique reads numbers of positive samples are 0.036M, 0.53M, and 0.24M, respectively. Sequencing depths are 40x, 660x, and 300x, respectively).

## FUTURE PERSEPCTIVE

Based on single molecule SBS technique, GenoCare™ 1600 sequencer is designed and developed for clinical applications. With simple library preparation, minimum facility requirement, and high sensitivity, GenoCare™ 1600 shows good performance in reproductive health field, such as NIPT, PGT and chromosome analysis of the miscarriage tissue [25, 26]. For many NGS-based clinical tests inside hospital, high cost per run, long hands-on operation time and complicated procedure would severely hinder adoption; while GenoCare™ 1600 offers improvements in all above aspects. In addition, the design of 16 physically divided lanes and separated loading fluidics reduces sample mixing for each run and allows sequencing of flexible numbers of samples. Increasing the numbers of test samples beyond 16 is also feasible, as we designed two types of index that enables sequencing maximum of 32 samples per run.

As discussed in this paper, GenoCare™ 1600 shows great potential in pathogenesis infection diagnostics. Aiming at different clinical departments’ requirements, we could build up auxiliary diagnosis system, which combines immunoassay, culturing technique, and clinical manifestations, etc. with sequencing, to quickly confirm the types of infections and aid effective treatment. In addition, screening and monitoring the variants of microorganism’s genome would help public-health professionals evaluate and predict the evolution of disease and optimize pandemic-control solutions. To achieve this goal, we will continue improving GenoCare™’s sequencing speed and accuracy. We are developing novel nucleic acid and enzyme chemistry to achieve four-nucleotide addition in each cycle to expedite sequencing. In addition, the Q-score, base calling algorithm, and bioinformatics software are being developed to reduce the error rate. We believe this platform could expedite the application of DNA sequencing in clinical diagnosis in the future.

## Data Availability

Supplementary material is submitted as "Supplementary Information-Two Color Single Molecule Sequencing on Genocare 1600 Platform to Facilitate Clinical Applications.pdf".

## ACKNOWLEDGEMENT

We would like to thank Shenzhen Uni-medica Co., Ltd who kindly provides us research samples.

We also thank all current and past members of the GeneMind team who contributed to the development of the sequencing technology.

## CONTRIBUTORS

FC, BL, MC, LL(Lu), QW, HS, BX, LL(Lei Liu), XL, WC, BY, DC, HL, NZ, BZ, WW, JC, SL, HD, XH, WWL, YL, JF and LS developed the sequencing chemistry (including virtual terminator, polymerase, surface chemistry, imaging buffer, etc.).

ZJ, ZZ, PW, JX, RS, GW, JZ (Zheng) and JQ developed GenoCare™ 1600 sequencing instrument.

HJ, LL(Li), WC, LZ and YZ developed basecalling, alignment, Q-score and bioinformatics algorithms.

MZ, GL, LL (Lichun Liu) and YL developed kits for library preparation, hybridization and coronavirus detection.

HS, HJ, JZ (Zhang), YF, JC and WH provided data analysis and experimental material support for this paper.

FC, LS, ZZ, MZ, and WP wrote the manuscript.

QY directed the project.

